# Timing of PCR and Antibody Testing in Patients with COVID-19 associated dermatologic manifestations

**DOI:** 10.1101/2020.07.03.20146134

**Authors:** Esther E. Freeman, Devon E. McMahon, Lindy P. Fox, Marlys S. Fassett

**Affiliations:** Department of Dermatology, Massachusetts General Hospital, Harvard Medical School, Boston, MA; Medical Practice Evaluation Center, Mongan Institute, Massachusetts General Hospital, Boston, MA; Department of Dermatology, University of California San Francisco, San Francisco, CA; Department of Immunology and Microbiology, University of California San Francisco, San Francisco, CA

**Keywords:** COVID-19, SARS-CoV-2, dermatology, public health, registry, pernio, chilblains

## Abstract

A recent study from Spain noted 40 patients with chilblain-like lesions in suspected COVID-19.^1^ None tested PCR positive for SARS-CoV-2, but 30% had detectable antibodies. The rapid increase in chilblain/pernio-like cases during the COVID-19 pandemic is likely SARS-CoV-2-associated. The relationship between skin symptom onset and COVID-19 PCR/antibody test timing, however, remains uncharacterized.

We established an international registry for cutaneous manifestations of COVID-19.^2, 3^ Providers reported time between dermatologic symptom onset and positive/negative COVID-19 laboratory results, when available.

From 8 April-30 June, 2020, 906 laboratory-confirmed or suspected COVID-19 cases with dermatologic manifestations were reported, 534 of which were chilblains/pernio.^3^ Among PCR-tested patients, 57%(n=208) overall and 15%(n=23) of chilblains/pernio cases were PCR-positive. Antibody positivity was 37%(n=39) overall and 19%(n=15) for chilblains/pernio.

We evaluated 163 patients with timing information on PCR and/or antibody testing (Table 1). For patients with suspected COVID-19 and any cutaneous manifestation, PCR-positive testing occurred median 6 (IQR 1-14) days after dermatologic symptoms started while PCR-negative testing occurred median 14 (IQR 7-24) days later. For patients with pernio/chilblains, PCR-positivity was noted 8 (IQR 5-14) days after symptoms and negativity median 14 (IQR 7-28) days later. Antibody testing (IgM or IgG) was positive median 30 (IQR 19-39) days after symptom onset for all dermatologic manifestations and 27 (IQR 24-33) days after chilblains/pernio onset.

Like Hubiche et al, our data highlight the low frequency of SARS-CoV-2 PCR+ testing in COVID-19 patients with cutaneous manifestations. Positive predictive values for COVID-19 PCR are influenced by viral shedding kinetics, which are difficult to assess in non-respiratory presentations.^4^ Our data reveal that early PCR testing is more likely to be positive than later testing, even when date-of-onset is defined by cutaneous manifestations rather than systemic symptoms.

Most COVID-19 antibody data are from systemically-ill patients; the kinetics of antibody production in mild-to-moderate COVID-19 infections remain unclear.^5^ Here, positive antibodies resulted median 30 days from disease onset, beyond the frequently used 14-21 day testing window. In outpatients with true infection, many factors influence the likelihood of a positive antibody result: antibody production, test availability, assay sensitivity, and timing of care-seeking in relation to symptom-onset. These variables influence our interpretation of individual test results and our understanding of the association between pernio and COVID-19.

More population-level testing data is necessary to optimize diagnostic test timing. Positive identification of COVID-19 in minimally-symptomatic patients, including patients with skin findings, is critical to the public health effort.

## Manuscript Text

A recent study from France noted 40 patients with chilblain-like lesions in suspected COVID-19.^1^ None tested PCR positive for SARS-CoV-2, but 30% had detectable antibodies. The rapid increase in chilblain/pernio- like cases during the COVID-19 pandemic is likely SARS-CoV-2-associated. The relationship between skin symptom onset and COVID-19 PCR/antibody test timing, however, remains uncharacterized.

**Table 1:**
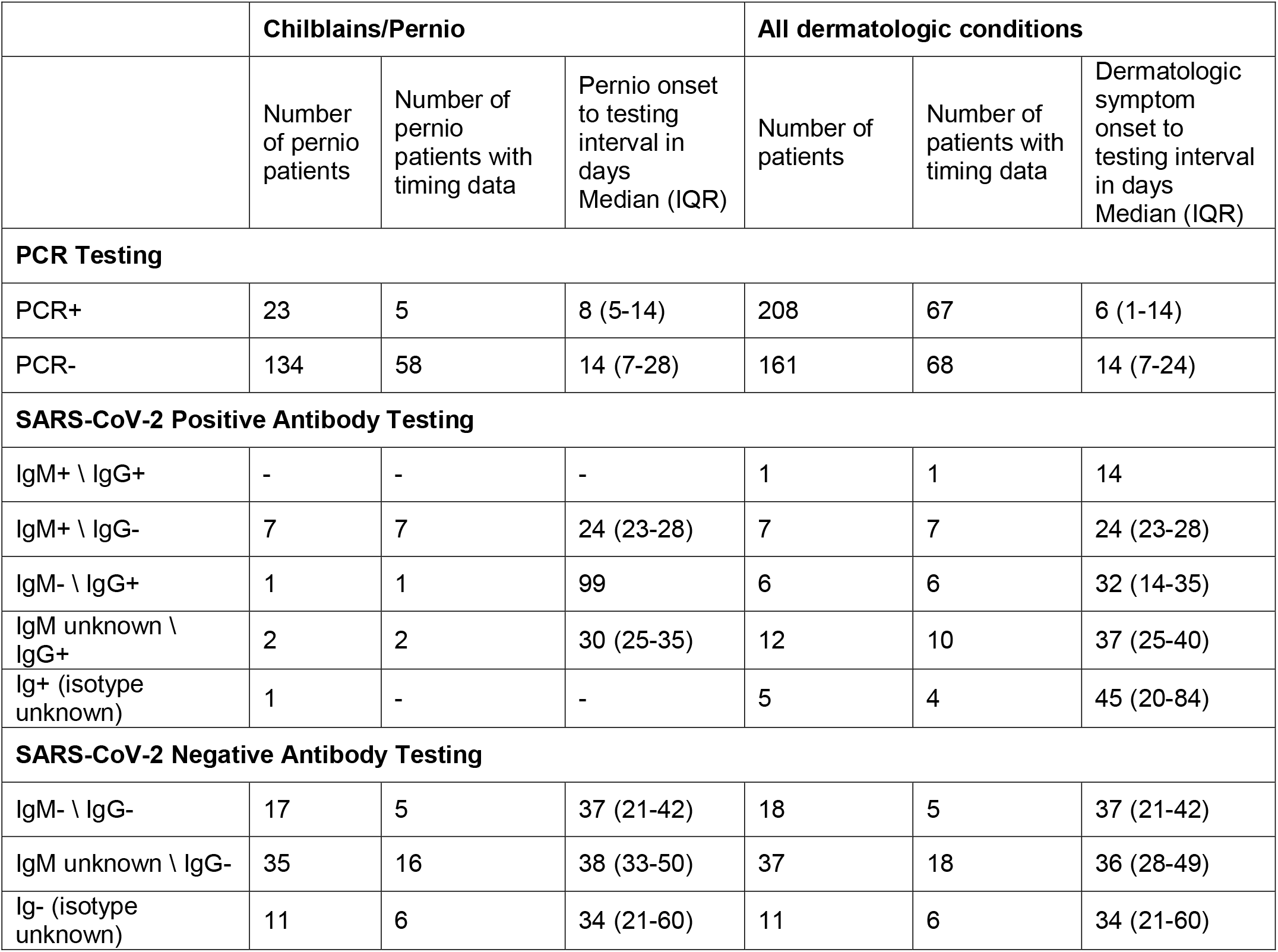
Distribution and timing of SARS-CoV-2 PCR and COVID-19 antibody test results in relation to dermatologic manifestations.

Like Hubiche *et al*, our data highlight the low frequency of SARS-CoV-2 PCR+ testing in COVID-19 patients with cutaneous manifestations. Positive predictive values for COVID-19 PCR are influenced by viral shedding kinetics, which are difficult to assess in non-respiratory presentations.^4^ Our data reveal that early PCR testing is more likely to be positive than later testing, even when date-of-onset is defined by cutaneous manifestations rather than systemic symptoms.

## Data Availability

Data is not available.

## Ethics

The registry was reviewed by the Partners Healthcare (MGH) Institutional Review Board (IRB) and was determined to not meet the definition of Human Subjects Research.

